# A SARS-CoV-2 lineage A variant (A.23.1) with altered spike has emerged and is dominating the current Uganda epidemic

**DOI:** 10.1101/2021.02.08.21251393

**Authors:** Daniel Lule Bugembe, My V.T.Phan, Isaac Ssewanyana, Patrick Semanda, Hellen Nansumba, Beatrice Dhaala, Susan Nabadda, Áine Niamh O’Toole, Andrew Rambaut, Pontiano Kaleebu, Matthew Cotten

## Abstract

SARS-CoV-2 genomic surveillance in Uganda provides an opportunity to provide a focused description of the virus evolution in a small landlocked East African country. Here we show a recent shift in the local epidemic with a newly emerging lineage A.23 evolving into A.23.1 which is now dominating the Uganda cases and has spread to 26 other countries. Although the precise changes in A.23.1 as it has adapted are different from the changes in the variants of concern (VOC), the evolution shows convergence on a similar set of proteins. The A.23.1 spike protein coding region has accumulated changes that resemble many of the changes seen in VOC including a change at position 613, a change in the furin cleavage site that extends the basic amino acid motif, and multiple changes in the immunogenic N-terminal domain. In addition, the A.23.1lineage encodes changes in non-spike proteins that other VOC show (nsp6, ORF8 and ORF9). The clinical impact of the A.23.1 variant is not yet clear, however it is essential to continue careful monitoring of this variant, as well as rapid assessment of the consequences of the spike protein changes for vaccine efficacy.

## Main Text

The novel Severe Acute Respiratory Syndrome Coronavirus 2 (SARS-CoV-2)(1) and the associated disease Coronavirus Disease 2019 (COVID-19)(2)(3) continue to spread throughout the world, causing >120 million infections and >2.6 million deaths (16 Mar 2021, <u>Johns Hopkins COVID-19 Dashboard</u>). Genomic surveillance has played a key role in the response to the pandemic; sequence data from SARS-CoV-2 provides information on the transmission patterns and the evolution of the virus as it enters new regions and spreads. As COVID-19 vaccines become available and are implemented, monitoring SARS-CoV-2 genetic changes, especially changes at epitopes with implications for immune escape is crucial. A detailed classification system has been defined to help monitor SARS-CoV-2 as it evolves (4) with virus sequences classified into 2 main phylogenetic lineages (Pango lineages) A and B, representing the earliest divergence of SARS-CoV-2 in the pandemic and then into sub-lineages within these. Several Variants of Concern (VOC) have emerged showing increased transmission patterns and reduced susceptibility to vaccine and/or therapeutic antibody treatments. These VOC include lineage B.1.1.7 first identified in the UK (5), B.1.351 in South Africa (6) and lineage P.1 (B.1.1.28.1) in Brazil (7).

### Status of the SARS-CoV-2 epidemic in Uganda

SARS-CoV-2 infection was first detected in Uganda in March 2020, initially among international travellers until passenger flights were stopped in late March 2020. A second route of virus entry with truck drivers from adjacent countries then became apparent (8). Since August 2020, community transmission dominated the Uganda case numbers. By March 2021 total cases in Uganda were 40,535, with 334 deaths attributed to the virus. We have continued our efforts to generate SARS-CoV-2 genomic sequence data to monitor virus movement and genetic changes and we report here on a novel sub-lineage A (A.23.1) that emerged and is dominating the local epidemic. The A.23.1 variant encodes multiple changes in the spike protein as well as in nsp6, ORF8 and ORF9, some predicted to be functionally similar to those observed in VOC in lineage B.

### Changes in prevalence of lineage A viruses

The genomes generated here were classified into Pango lineages(4) using the Pangolin module pangoLEARN (https://github.com/cov-lineages/pangolin) and into NextStrain clades using NextClade (9) (https://clades.nextstrain.org/). The distribution of virus lineages circulating in Uganda changed dramatically over the course of the year. A clear feature of the earlier COVID-19 epidemic in the country was the diversity of viruses found throughout the country attributed to frequent flights into Uganda from Europe, UK, US and Asia; this is reflected in the 9 lineages seen from March to May 2020 with a mixture of both lineage A and B viruses (Figure 1, panel a). After passenger flights were limited in March 2020, the virus entered via land travel with truck drivers. Uganda is landlocked country, characterised by its important geographical position, i.e. the crossing of two main routes of the Trans-Africa Highway in East Africa. The essential nature of produce and goods transport allowed virus movement from/to Kenya, South Sudan, DRC, Rwanda and Tanzania. In the period of June to August 2020 lineage B.1 and B.1.393 strains were abundant, similar to patterns observed in Kenya (10) (Figure 1, panel b) although lineage A viruses did not decline as seen in US and Europe. Lineage A.23 strains were first observed in two prison outbreaks in Amuru and Kitgum, Uganda in August 2020 and by the September-November period, the A.23 was the major lineage circulating throughout the country (Figure 1, panel c). The A.23 virus continued to evolve into the lineage A.23.1, first observed in late October 2020. Given the diversity of virus lineages found in the country from March until November 2020, it was unexpected that by late December 2020 to January 2021, lineage A.23.1 viruses represented 90% (102 of 113 genomes) of all viruses observed in Uganda (Figure 1, panel d). In all time periods, the SARS-CoV-2 positive sample were obtained from multiple clinical and surveillance locations throughout Uganda indicating that the differences are unlikely to be due to sampling different subpopulations in the country at different times.

**Figure 1.**
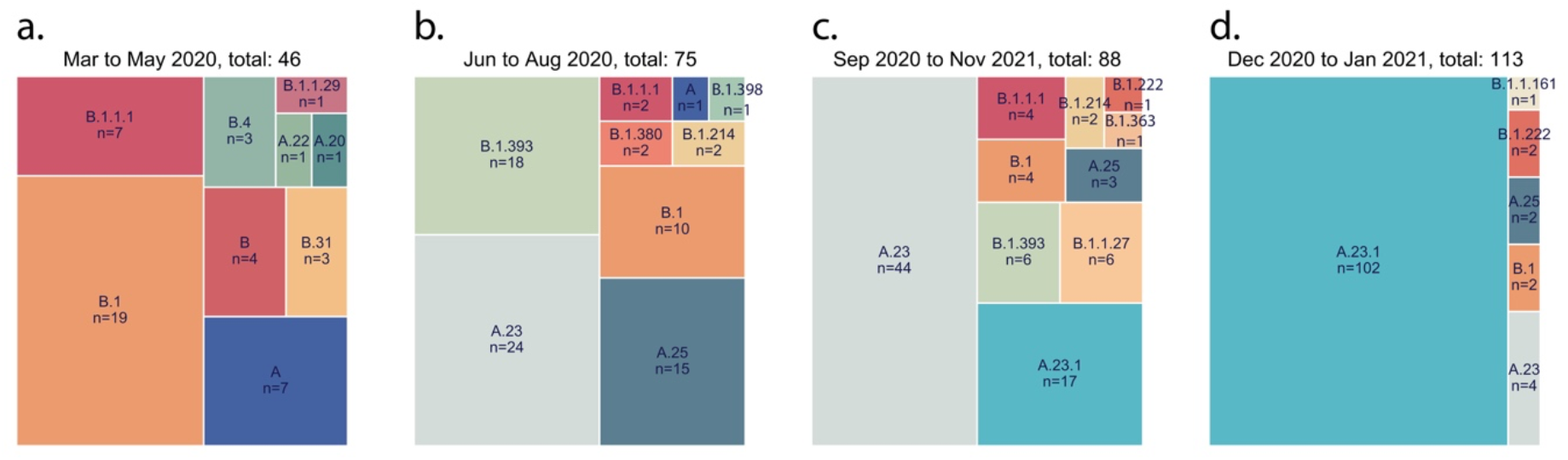
SARS-CoV-2 lineage diversity in Uganda. All high-coverage complete from Uganda (n=322) were lineage typed using the pangolin resource (https://github.com/cov-lineages/pangolin) Lineage counts were stratified into four periods (**panel a:** March-May 2020, **panel b:**June-August 2020, **panel c:** September to November 2020 and **panel d:** December 2020 to January 2021). The percentage of each lineage within each set was plotted as a treemap using squarify (https://github.com/laserson/squarify) with the size of each sector proportional to the number of genomes, genomes numbers are listed with “n=”.

### Virus sequence diversity including fatal cases

All newly and previously generated Uganda genomes that were complete and high-coverage (n=322) were used to construct a maximum-likelihood phylogenetic tree (Figure 2).

**Figure 2.**
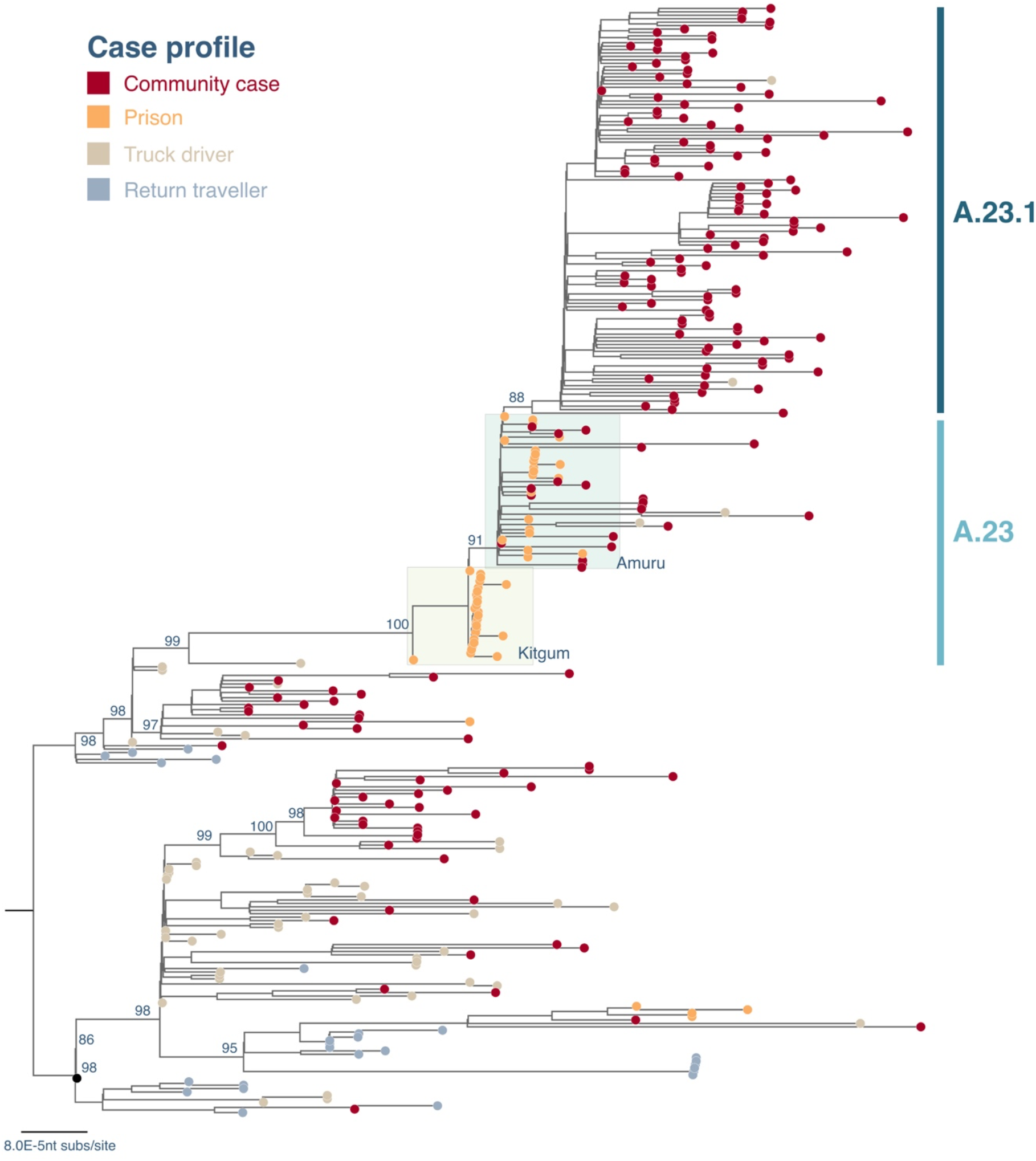
Maximum-likelihood phylogenetic tree comparing all available complete and high-coverage Uganda sequences (N=322). Strain names are coloured according to the case profile (cases from the community: dark red, prison: orange, truck driver: light brown, return traveller: light blue). The case clusters from prisons in Kitgum and Amuru are highlighted in colour boxes in light yellow and light green, respectively. The lineages A.23 and A.23.1 are indicated. The tree was rooted where lineages A and B were split. The branch length is drawn to the scale of number of nucleotide substitutions per site, indicated in lower left, and only bootstrap values of major nodes were shown.

A number of A and B variant lineages were observed briefly at low frequencies and may have undergone extinction, similar to patterns observed in the UK (11) and Scotland (12). Genomes identified from a truck driver are often observed basal to community clusters (Figure 2), suggesting the importance of this route in the introduction and spread of the virus into Uganda. Most of genomes from truck drivers sampled at points of entry (POEs) bordering Kenya belonged to lineage B.1 and B.1.393 consistent with the pattern reported in Kenya (10). However, genomes identified from truck drivers from Tanzania, and from the Elegu POE bordering South Sudan, albeit small numbers, belonged to both A and B.1 lineages. Continued monitoring of truck drivers coming in and out of the Uganda provides a useful description of the inland circulation of strains in this part of world, where genomic surveillance is not as detailed as in other parts of the world.

### Emergence of A.23 and A.23.1

Outbreaks of SARS-CoV-2 infections were reported in the Amuru and Kitgum prisons in August 2020 (13)(14). The SARS-CoV-2 genome sequences from individuals in the prisons were exclusively belonging to lineage A (Figure 2) with three amino acid (aa) changes encoded in the spike protein (F157L, V367F and Q613H, Figure 3) that now define lineage

**Figure 3.**
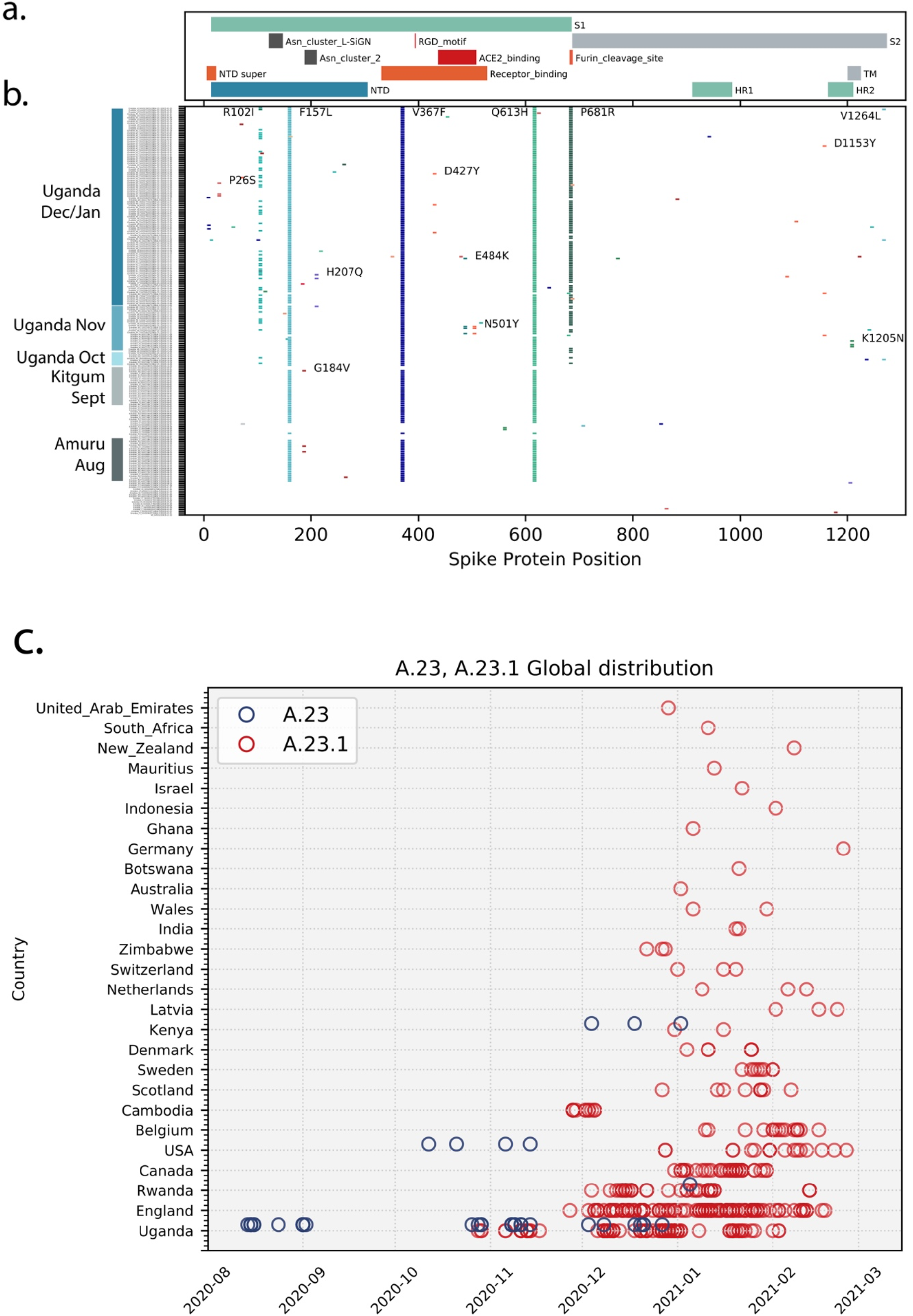
Spike protein changes in lineage A.23 and A.23.1 relative to the SARS-CoV-2 reference strain (NC_045512) encoded protein are documented. **Panel a**: The locations of important spike protein features are indicated. NTD: N-terminal domain, RBD: receptor-binding domain, S1: spike 1, S1: Spike 2, TM: transmembrane domain, HR1: helical repeat 1, HR2: helical repeat 2, NTD super: N-terminal domain supersite. **Panel b:** Each line represents the encoded spike protein sequence from a single genome, ordered by date of samples collection (bottom earliest, top most recent). Sequences from Amuru in August 2020, Kitgum in September 2020 and Uganda in October, November, December 2020/January 2021 are indicated. Markers indicating the positions of amino acid (aa) differences from the reference strain, changes observed in multiple genomes are annotated with the annotation (original aa position new aa). **Panel c. Current global distribution of and A.23.1** All available SARS-CoV-2 complete genomes annotated as complete and lineage A from GISAID were retrieved on Feb 4 2021 and lineage typed using Pangolin(19). and confirmed as A.23 and A.23.1 by extracting examining the encoded spike protein. All novel Uganda A.23 and A.23.1 reported here were also included. Genomes were plotted by country and sample collection date.

A.23 (see below). By October 2020, lineage A.23 viruses were also found outside of the prisons in a community sample from Lira (a town 140 km from Amuru), in two samples from the Kitgum hospital, in several community samples from Kampala, Jinja, Mulago, Tororo, Soroti as well as in 2 truck drivers collected at POE bordering Kenya. By November 2020, the A.23 viruses spread further to northern Uganda in Gulu and Adjumani. Lineage A.23 viruses were not seen in Uganda (or anywhere in the world) before August 2020 (Figure 3 panel c), yet the A.23 viruses were attributed to 32% of the viruses in Uganda (Figure 1) from June to August 2020 and 50% of the observed viruses in September to November 2020. In late October, the A.23.1, a variant evolving from A.23, with additional change in the spike protein (P681R) was observed (Figure 3, panel b, c) and by December 2020-January 2021, 90% of identified genomes (102 out of 113) belonged to the new A.23.1 lineage (Figure 1 and 2). The mutations in A.23.1 were consistent with evolution from an original A.23 virus observed in Amuru/Kitgum cluster (Figure 2 and Supplemental Figure 1) as well as changes nsp6 and ORF9 (Supplemental Figure 2 and 4).

### Important changes observed in the spike protein

The spike protein is crucial for virus entry into host cells, for tropism, and is a critical component of COVID-19 vaccine development and monitoring. The changes in spike protein observed in Uganda and global A.23 and A.23.1 viruses are shown in Figure 3 panel b.

Many amino acid (aa) changes were single events with no apparent transmission observed. However, the initial lineage A.23 genomes from Amuru and Kitgum encoded three amino acid changes in the exposed S1 domain of spike (F157L, V367F and Q613H, Figure 3 panel b). The V367F change is reported to modestly increase infectivity(15), the Q613H change may have similar consequences as the D614G change observed in the B.1 lineage found predominantly in Europe and USA; in particular, D614G was reported to increase infectivity, spike trimer stability and furin cleavage (15),(16),(17),(18). These changes were not observed in previously reported genomes from Uganda (8). Of some concern, the mutations E484K and N501Y amino acid changes in the receptor binding domain (RBD) were observed in the A.23 viruses identified in Adjumani cases on 9^th^ to 11^th^ November 2020 (Figure 3, panel b). These two amino acid changes are shown to substantially compromise the vaccine efficacy as well as antibody treatments.

Of concern, the recent Kampala and global A.23.1 virus sequences from December 2020-January 2021 now encoded 4 or 5 amino acid changes in the spike protein (now defining lineage A.23.1, see below) plus additional protein changes in nsp3, nsp6, ORF8 and ORF9 (Figure 3 panel b, Figure 4). The P681R spike change adds a basic amino acid adjacent to the spike furin cleavage site. This same change has been shown *in vitro* to enhance the fusion activity of the SARS-CoV-2 spike protein, likely due to increased cleavage by the cellular furin protease (20); importantly, a similar change (P681H) is encoded by the recently emerging VOC B.1.1.7 that is now spreading globally across 75 countries as of 5 February 2021 (5) (21). There are also changes in the spike N-terminal domain (NTD), a known target of immune selection, observed in samples from Kampala A.23.1 lineage, including P26S and R102I (Figure 3 panel b). Additionally and importantly, a A.23.1 strain identified in Kampala on 11^th^ December 2020 carried the E484K change in the RBD, which may add further concern of this particular variant as it gains higher transmissibility and enhanced resistance to vaccine and therapeutics. Outside of the spike protein, a single nucleotide change (G27870T) leading to early termination of the ORF7b (E39*) was observed in the A.23.1 from the community cases in Tororo in late December 2020. Although the clinical implication of this change is yet to be determined, it is important to document such change for further follow-up.

**Figure 4.**
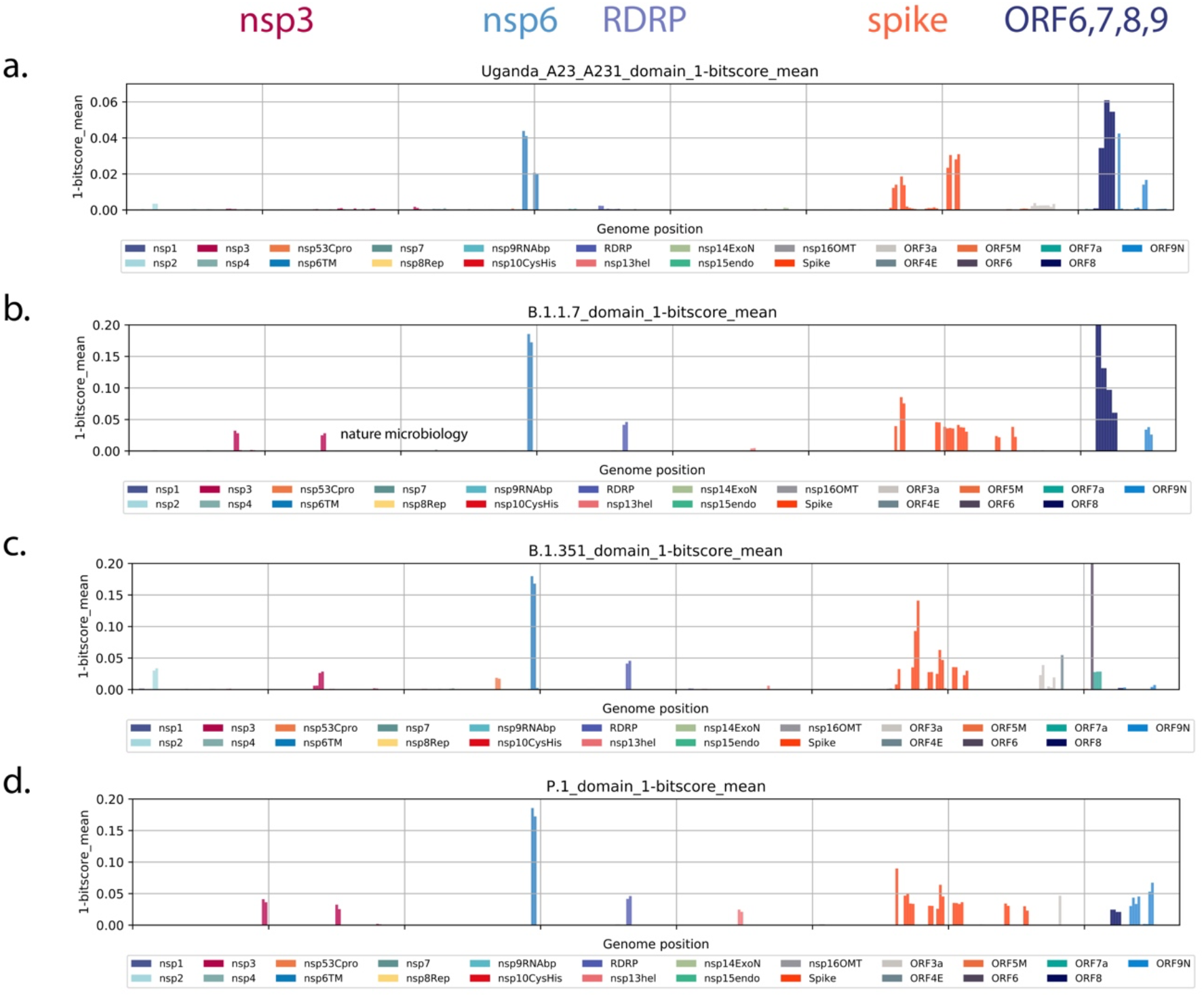
All protein changes across lineage new variants. All forward open reading frames from the 35 early lineage B SARS-CoV-2 genomes were translated, and processed into 44 aa peptides (with 22 aa overlap), clustered at 0.65 identity using uclust (25), aligned with MAAFT (26) and converted into pHMMs using HMMER-3 (27). The presence of each domains and its bit-score (a measure of the similarity between the query sequence and the sequences used for the pHMM(27)) was sought in each set of SARS-CoV-2 VOC genomes and the 1-mean of the normalized domain bit-scores was plotted across the genome (e.g. 1 - the similarity of the identified query domain to the reference lineage B SARS-CoV-2 domain). Domains were coloured by the proteins from which they were derived with the colour code indicated below the figure. **Panel a**. Query set are 49 most Uganda lineage genomes. **Panel b**. All B.1.1.7 full genomes lacking Ns deposited in GISAID on Jan 26 2021, **Panel c**. All B.1.351 full genomes lacking Ns present in GISAID on Jan 26 2021, **Panel d**. All P.1 full genomes lacking Ns present in GISAID on Jan 26 2021.

### New lineage A designations

The viruses detected in Amuru and Kitgum met the criteria for a new SARS-CoV-2 lineage (22)(23) by clustering together on a global phylogenetic tree, sharing epidemiological history and source from a single geographical origin, and encoding multiple defining SNPs. These features including especially the three spike changes F157L, Q613H and V367F define the new lineage A.23. Continued circulation and evolution of A.23 in Uganda was observed and two additional changes in spike R102I and P681R were observed in December 2020 in Kampala; these SNPs define the sub-lineage A.23.1. Additional changes in non-spike regions also define the A.23 and A.23.1, including nsp3: E95K, nsp6: M86I, L98F, ORF 8: L84S, E92K and ORF9 N: S202N, Q418H. These new lineages can be assigned since pangolin version v2.1.10 and pangoLEARN data release 2021-02-01.

Screening SARS-CoV-2 genomic data from GISAID (March 12, 2021), A.23 and A.23.1 viruses are now found in 26 countries outside of Uganda (Figure 3 panel c). The A.23 was first observed in Uganda in August 2020, subsequently in USA in October and Kenya and Rwanda in December (Figure 3 panel c). The A.23.1 was first seen in Uganda in the community cases in Mbale on 28^th^ October 2020 and in Jinja on 29^th^ October 2020, and soon spreading across the country in early November 2020. Outside of Uganda, the A.23.1 was found in England and Cambodia from the end of November, in Rwanda from the beginning of December. Of note, the international flights out of Uganda were restarted on 1 October 2020 with flights to Europe, Asia and USA. Phylogenetic analysis supports the close evolution of A.23 to A.23.1 (Supplementary Figure 1).

### Additional changes in Ugandan A.23 and A.23.1 genomes compared to other VOC genomes

Although a main focus has been on spike protein changes, there are changes in other genomic regions of the SARS-CoV-2 virus accompanying the adaptation to human infection. We employed profile Hidden Markov Models (pHMMs) prepared from 44 amino acid peptides across the SARS-CoV-2 proteome (24) to detect and visualize protein changes from the early lineage B reference strain NC_045512. Measuring the identity score (bit-score) of each pHMMs across a query genome provides a measure of protein changes in 44 amino acid steps across the viral genome (Figure 4 panel a). This method applied to A.23 and A.23.1 genome sequences revealed the changes in spike (discussed above) as well as changes in the transmembrane protein nsp6 and the interferon modulators ORF8 and 9 (Figure 5 panel a).

We asked if a similar pattern of evolution was appearing in VOC as SARS-CoV-2 adapted to human infection. We gathered the sets of genomes described in the initial published descriptions of these VOC (B.1.1.7 (5), B.1.351 (28) or P.1 (7)) and applied the same profileHMM analysis. Similar to A.23/A.23.1, the B.1.1.7 lineage encodes nsp6, spike, ORF8 and 9 changes as well as changes in nsp3 and the RNA-dependent RNA polymerase (RRP, Figure 4 panel b). Lineage B.1.351 encodes nsp3, nsp6, RDRP, spike and ORF6 changes (Figure 4 panel c) and lineage P.1 encodes nsp3, nsp6, RDRP, nsp13, spike and ORF8 and 9 changes (Figure 4 panel d). Although the exact amino acid and positions of change within the proteins differ in each lineage, there are some striking similarities in the common proteins that have been altered. Of interest, the nsp6 change present in B.1.1.7, B.1.351 and P.1 is a 3 amino acid deletion (106, 107 and 108) in a protein loop of nsp6 predicted to be on exterior of the autophagy vesicles on which the protein accumulates (29).The three amino acid nsp6 changes of lineage A.23.1 are L98F in the same exterior loop region, and the M86l and M183I changes predicted to be in intramembrane regions but adjacent to where the protein exits the membrane (29), (Supplementary Figure 2). A compilation of the amino acid changes in A.23.1 and the VOC lineages is found in Supplementary Table 2 with proteins that are altered in all 4 lineages marked in red.

## Discussion

We report the emergence and spread of a new SARS-CoV-2 variant of the A lineage (A.23.1) with multiple protein changes throughout the viral genome. A similar phenomenon recently occurred with the B.1.1.7 lineage, detected first in the southeast of England (5) and now globally and with the B.1.351 lineage in South Africa (6), and P.1 lineage in Brazil (30) suggesting that local evolution (perhaps to avoid the initial population immune responses) and spread may be a common feature of SARS-CoV-2. Importantly, lineage A.23.1 shares many features found in the lineage B VOC including: alteration of key spike protein regions, especially ACE2 binding region which is exposed and immunogenic, the furin cleavage site and the 613/614 change that may increase spike multimer formation. The VOC and A.23.1 strains also encode changes in similar region of the nsp6 protein which may be important for altering cellular autophagy pathways that promote replication. Changes or disruption of ORF7,8 and 9 are also present in the VOC and A.23.1. The ORF8 changes or deletion probably indicates this protein is unnecessary for human replication, similar deletions accompanied SARS-CoV-2 adaption to humans(31),(32).

We suspect that emerging SARS-CoV-2 lineages may be adjusting to infection and replication in humans and it is notable that the VOCs and lineage A.23.1 share some common features in their evolution. The spike changes are best understood due to the massive global effort to define the receptor and develop vaccines against the infection. The analysis reported in Figure 4 reveals common functions of SARS-CoV-2 that have been altered in all four variants, especially nsp6 and the ORFs 8 and 9. The functional consequences of the additional non-spike changes warrant additional studies and the current analysis may focus efforts of the proteins that are commonly changed in the variant lineages. Finally the susceptibility of A.23.1 to vaccine immune responses is of great importance to determine as vaccines become available in this part of Africa.

## Methods

### Sample collection, whole genome MinION sequencing and genome assembly

Residual nucleic acid extract from SARS-CoV-2 RT-PCR positive samples were obtained from Central Public Health Laboratory (Kampala, Uganda). The nucleic acid was converted to cDNA and amplified using SARS-CoV specific 1500bp-amplicon spanning the entire genome as previously described(33).The resulting DNA amplicons were used to prepare sequencing libraries, barcoded individually and then pooled to sequence on MinION R.9.4.1 flowcells, following the standard manufacturer’s protocol.

The genome assemblies were performed as previously described (8). Briefly, reads from fast5 files were base-called and demultiplexed using Guppy 3.6 running on the UMIC HPC. Adapters and primers sequences were removed using Porechop (https://github.com/rrwick/Porechop) and the resulting reads were mapped to the reference genome Wuhan-1 (GenBank NC_045512.2) using minimap2(34) and consensus genomes were generated in Geneious (Biomatters Ltd). Genome polishing was performed in Medaka, and SNPs and mismatches were checked and resolved by consulting raw reads.

### Phylogenetic analyses

For the local Uganda virus comparison, all newly and previously generated genomes from Uganda (N=322) were aligned using MAFFT (26) and manually checked in AliView (35). The 5’ and 3’ untranslated regions (UTRs) were trimmed. Maximum-likelihood (ML) phylogenetic tree was constructed using RAxML-NG (36) under the GTR+I+G4 model as best-fitted substitution model according to Akaike Information Criterion (AIC) determined by ModelTest-NG (37) and run for 100 pseudo-replicates. Resulting tree was visualised in Figtree(38) and rooted at the point of splitting lineage And B.

For phylogenetic analyses of Uganda lineage A.23 and A.23.1 strains comparing to global A.23/A.23.1 strains, the global SARS-CoV-2 lineage A.23 (N=8) and A.23.1 (N=38) genomes were retrieved from GISAID on 12 March 2021. These global A.23/A.23.1 genomes combining with Ugandan A.23/A.23.1 genomes (N=191) were aligned using MAFFT and manually checked in AliView, followed by trimming 5’ and 3’ UTRs. The global and Ugandan A.23/A.23.1 genomes were used to construct a ML tree under the GTR+I+G4 model as best-fitted substitution model according to AIC determined by ModelTest-NG (37) and run for 100 pseudo-replicates using RAxML-NG. Resulting tree was visualised in Figtree and rooted using the A.23 lineage.

Profile Hidden Markov Model (profileHMM) domain analysis of A.23/A.23.1 and VOC genomes was performed as previously described (24) with some changes. A database of profileHMMs was generated from the first 65 lineage B SARS-CoV-2 genome sequences. All 3 forward open reading frames of each genome were translated computationally and then sliced into 44 amino acid segment with overlapping with 22 amino acids. All 44 amino acid query peptides were then clustered with uclust (25) and their original identity and coordinates determined by blastp search against a protein database made from the NC_045512 reference strain.

Query sets of genomes were processed to remove any genomes containing Ns (which disrupt the HMM scoring process). The hmmscan function from HMMER-3 (27) was used with the early B database. Query matches were identified using an E-value cutoff of 0.0001 and the bit-score values for each hit (a measure of the distance between the query 44 amino acid peptide and the B-lineage reference) was collected. Bit-scores for each domain were normalized by dividing each query score by the maximum score for that domain (x/x_max). In all analyses the original B lineage NC_045512 reference genome was included to define the maximum bit-score.

## Ethical approvals

This study was approved by the Uganda Virus Research Institute-Research and Ethics Committee (UVRI-REC Federalwide Assurance [FWA] FWA No. 00001354, study reference. GC/127/20/04/771) and by the Uganda National Council for Science and Technology, reference number HS936ES. The novel reported SARS-CoV-2 genomes are available on GISAID (https://www.gisaid.org/) under the accession numbers EPI_ISL_954226-EPI_ISL_954300 and EPI_ISL_955136. A second tranche of genomes has been submitted and is awaiting accession numbers.

## Data Availability

The novel reported SARS-CoV-2 genomes are available on GISAID (https://www.gisaid.org/) under the accession numbers EPI_ISL_954226-EPI_ISL_954300 and EPI_ISL_955136.

## Acknowledgements

We thank all global SARS-CoV-2 sequencing groups for their open and rapid sharing of sequence data and GISAID for providing an effective platform for making these data available. We are grateful to the Oxford Nanopore Technologies and the ARTIC Network for their support and we thank Pope Moseley for his constructive comments on the manuscript. The SARS-CoV2 diagnostic and sequencing award is jointly funded by the UK Medical Research Council (MRC/UKRI) and the UK Department for International Development (DFID) under the MRC/DFID Concordat agreement (grant agreement number NC_PC_19060) and is also part of the EDCTP2 programme supported by the European Union. The UMIC high performance computer was supported by MRC (grant number MC_EX_MR/L016273/1) to PK. A.R. acknowledges the support of the Wellcome Trust (Collaborators Award 206298/Z/17/Z <u>ARTIC network</u>) and the European Research Council (grant agreement no. 725422 – ReservoirDOCS). The study is additionally funded by the Wellcome, DFID - Wellcome Epidemic Preparedness – Coronavirus (grant agreement number 220977/Z/20/Z) awarded to MC.

## Author contribution statement

All authors contributed to the work presented in this paper.

## Competing Interests statement

The authors declare no competing interests.

## Supplementary Information

**Supplemental Figure 1.**
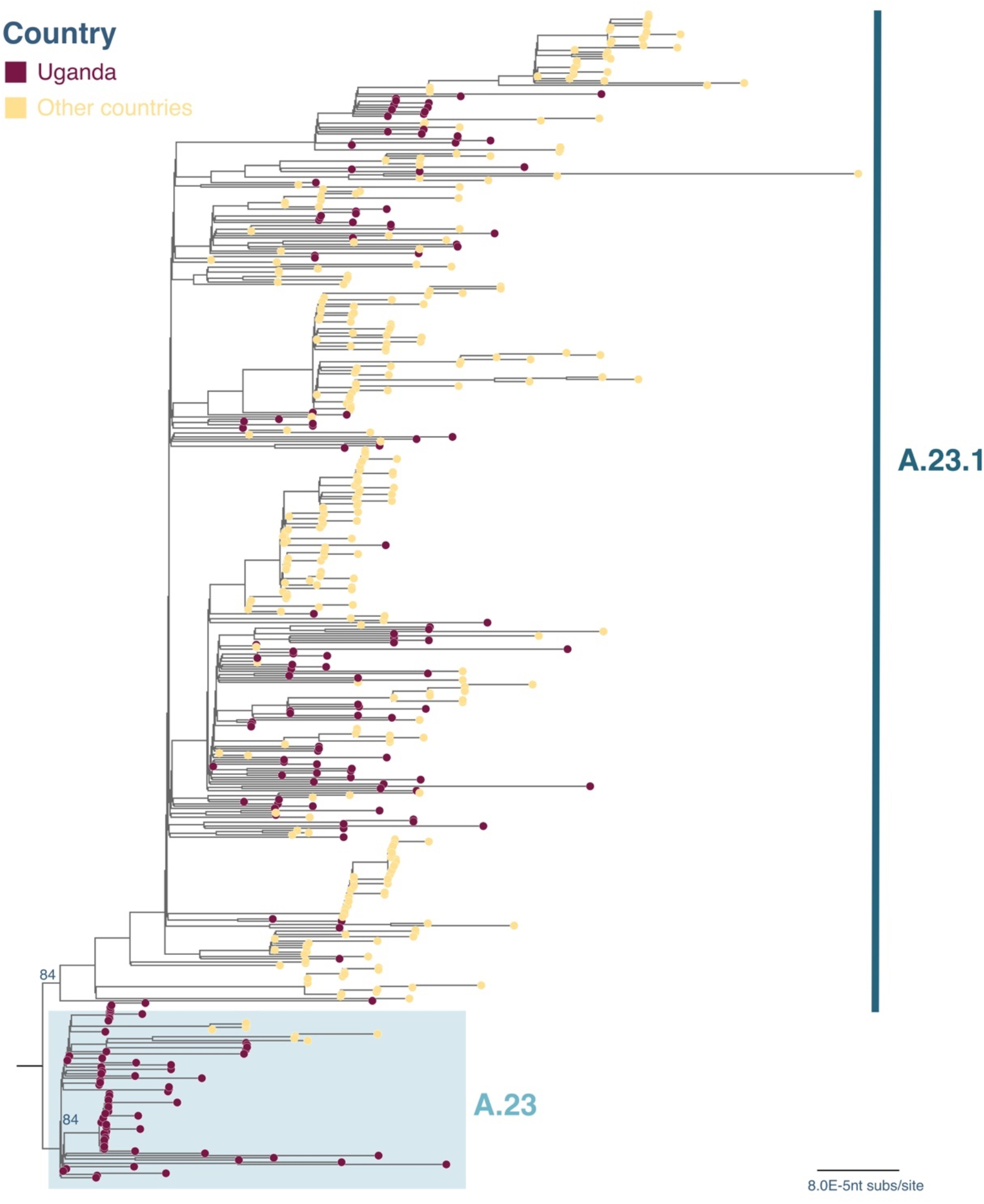
Maximum-likelihood phylogenetic tree comparing Uganda lineage A.23 and A.23.1 strains to global lineage A.23 and A.23.1 genomes. A maximum-likelihood (ML) phylogenetic tree comparing Ugandan A.23 and A.23.1 (N=191) with the global A.23 and A.23.1 (N=336). The tree was rooted by the A.23 lineage and strains were coloured according to the countries where they were identified. Branch length was drawn to the scale of number of nucleotide substitutions per site and only bootstrap values at the major nodes were shown. The tree was visualised in Figtree (38).

**Supplementary Table 1.** Lineage distribution in Uganda.

| Lineage | Count in Uganda<sup>1</sup> |  |
| --- | --- | --- |
| A | 8 |  |
| A.20 | 1 |  |
| A.22 | 1 |  |
| A.23 | 72 |  |
| A.23.1 | 119 |  |
| A.25 | 20 | A: 221 (69%) |
| B | 4 |  |
| B.1 | 35 |  |
| B.1.1.1 | 13 |  |
| B.1.1.161 | 1 |  |
| B.1.1.27 | 6 |  |
| B.1.1.29 | 1 |  |
| B.1.214 | 4 |  |
| B.1.222 | 3 |  |
| B.1.363 | 1 |  |
| B.1.380 | 2 |  |
| B.1.393 | 24 |  |
| B.1.398 | 1 |  |
| B.31 | 3 |  |
| B.4 | 3 | B: 101(31%) |
| Total | 322 |  |

1. All available genomes from Uganda were classified into Pango lineages using pangolin (19).

## Supplementary Information

**Supplementary Table 2.**
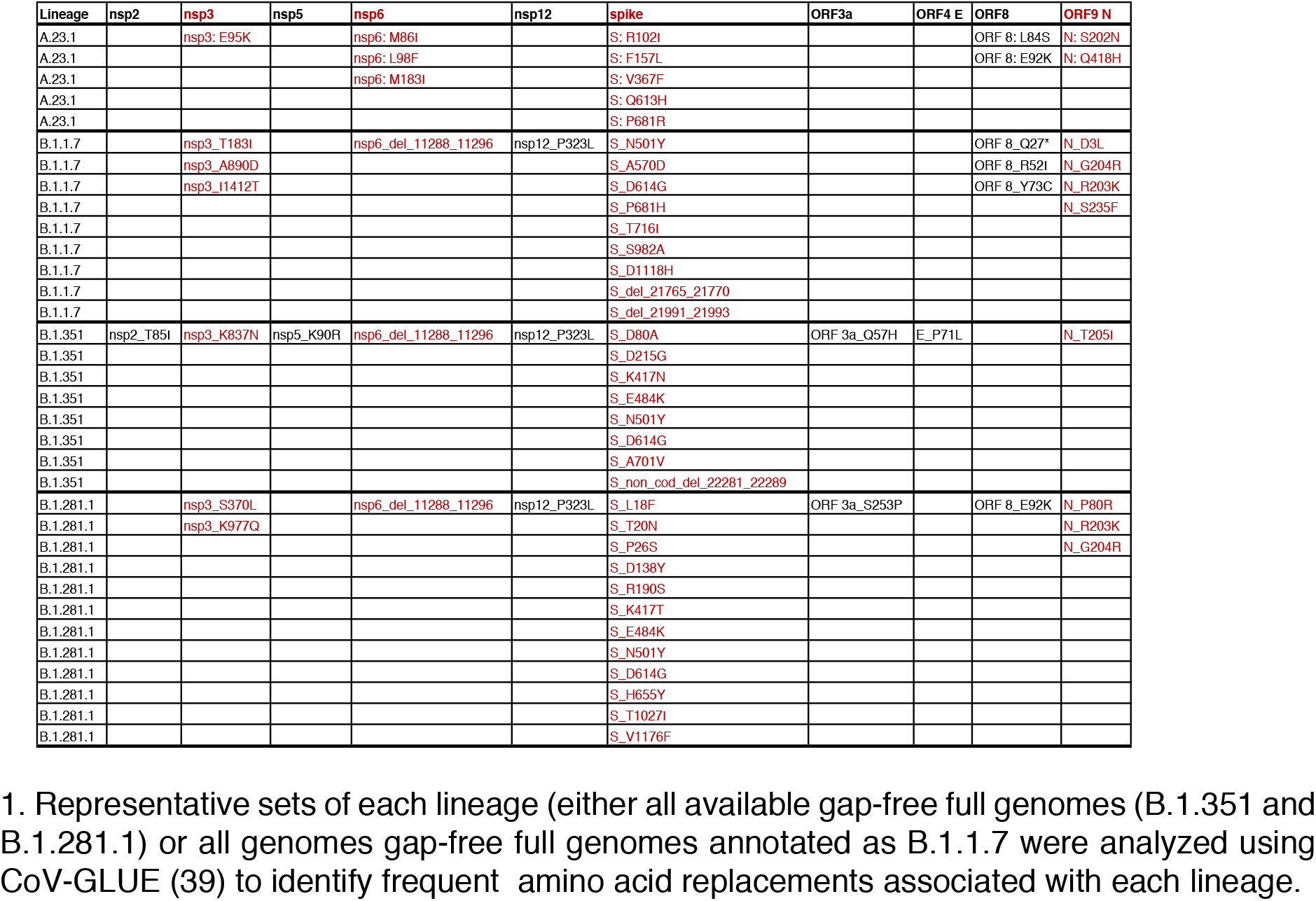
Summary of replacements in the A.23.1 and 3 VOC lineages^1^.

**Supplementary Figure 2.**
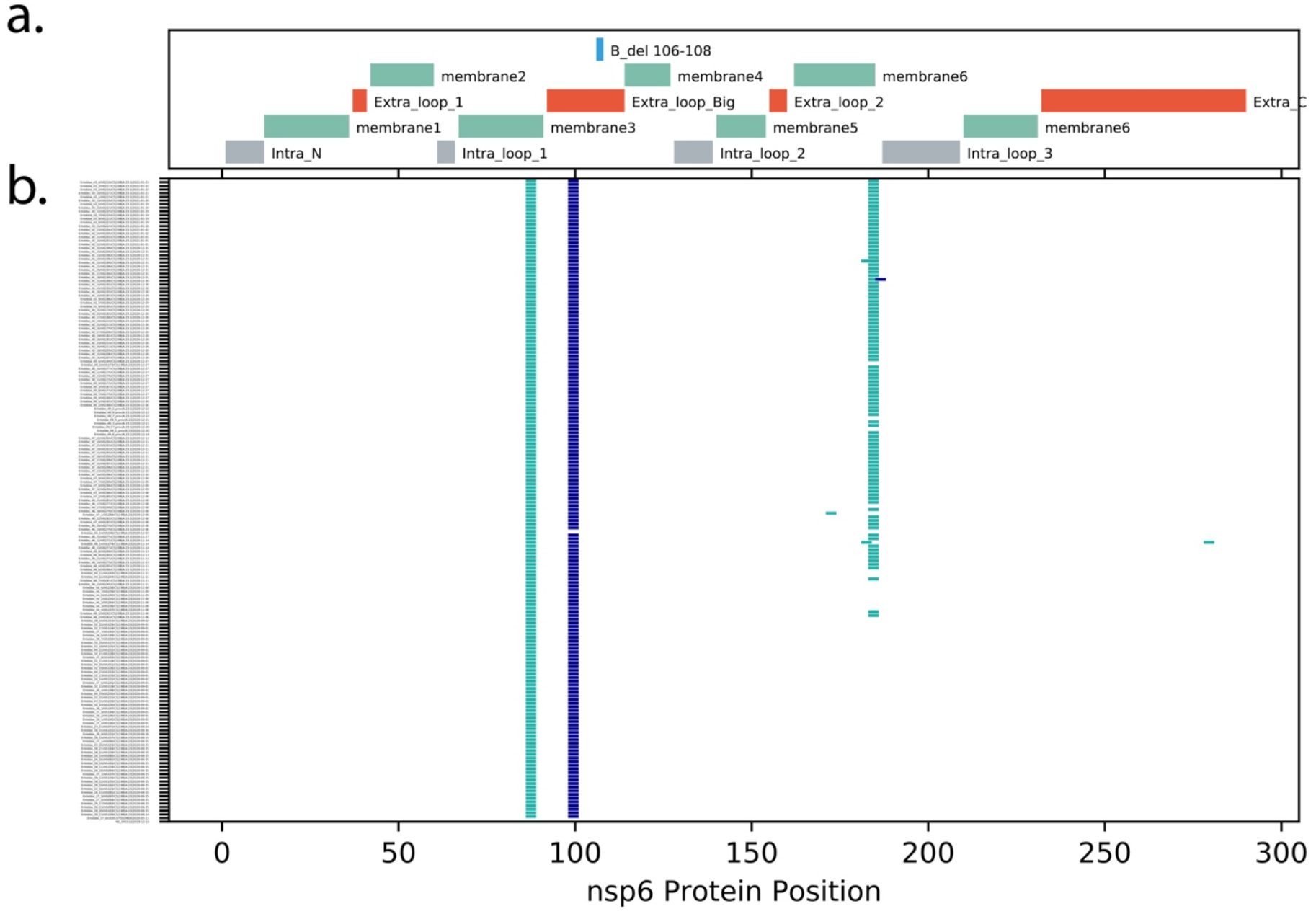
Changes in A.23/A.23.1 nsp6 protein. The encoded nsp6 protein from all Ugandan A.23 and A.23.1 genomes gather, aligned and compared to the nsp6 protein from GenBank NC_045512.2. **Panel a**: The locations of important nsp6 protein features are indicated based on the analysis of nsp6 from Benvenuto et al. (29). Intra_N: intravesicular amino-terminal region, Extra_loop_1: extravesicular loop1, Intra_loop_1: intravesicular loop 1, B_del 106-108: the region of nsp6 deleted in the lineage B VOC genomes, Extra_loop_Big: large extravesicular loop, Intra_loop_2: intravesicular loop 2, Extra_loop_2: extravesicular loop 2, Intra_loop_3: intravesicular loop 3, Extra_C: carboxy-terminal extra-vesicular portion. All features with “membrane” indicate membrane-spanning regions of nsp6. **Panel b:** Each line represents the encoded nsp6 protein sequence from a single genome, ordered by date of samples collection (bottom earliest, top most recent). Markers indicating the positions of amino acid (aa) differences from the reference strain, changes observed in multiple genomes are annotated with the annotation (original aa position new aa).

**Supplementary Figure 3.**
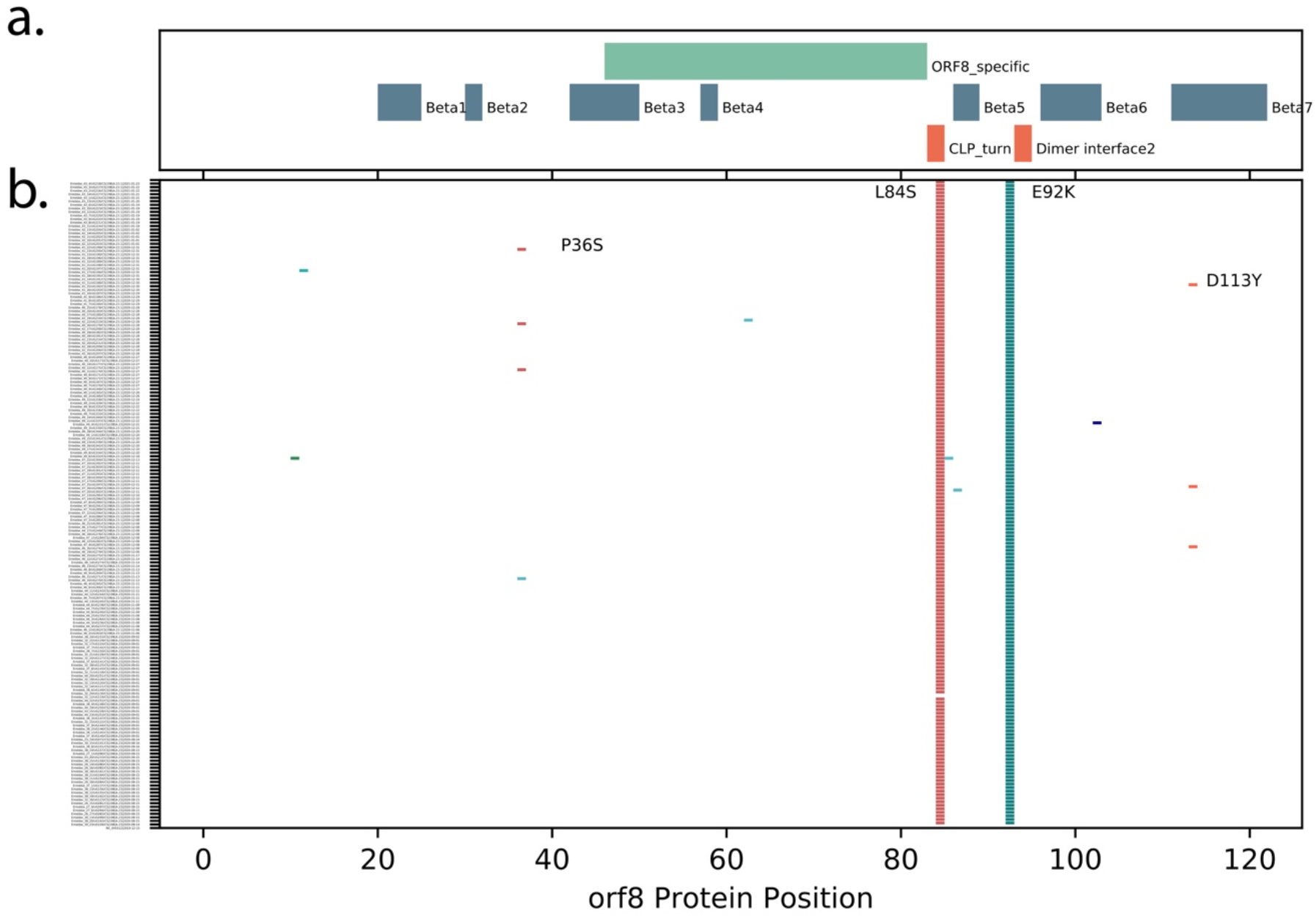
Changes in A.23/A.23.1 ORF8 protein. The encoded ORF8 protein from all Ugandan A.23 and A.23.1 genomes gather, aligned and compared to the ORF8 protein from GenBank NC_045512.2. **Panel a**: The locations of important ORF8 protein features are indicated based on the analysis of ORF8 from Flower et al. (40). Features with “Beta” indicate beta-sheets, ORF8_specific is a region unique to SARS-CoV-2 ORF8, CLP_turn: indicates a cysteine, Leucine, Proline motif essential for a fold in the mature protein, Dimer interface2 indicates the region of the protein the forms the interface between two monomers. **Panel b:** Each line represents the encoded ORF8 protein sequence from a single genome, ordered by date of samples collection (bottom earliest, top most recent). Markers indicating the positions of amino acid (aa) differences from the reference strain, changes observed in multiple genomes are annotated with the annotation (original aa position new aa).

**Supplementary Figure 4.**
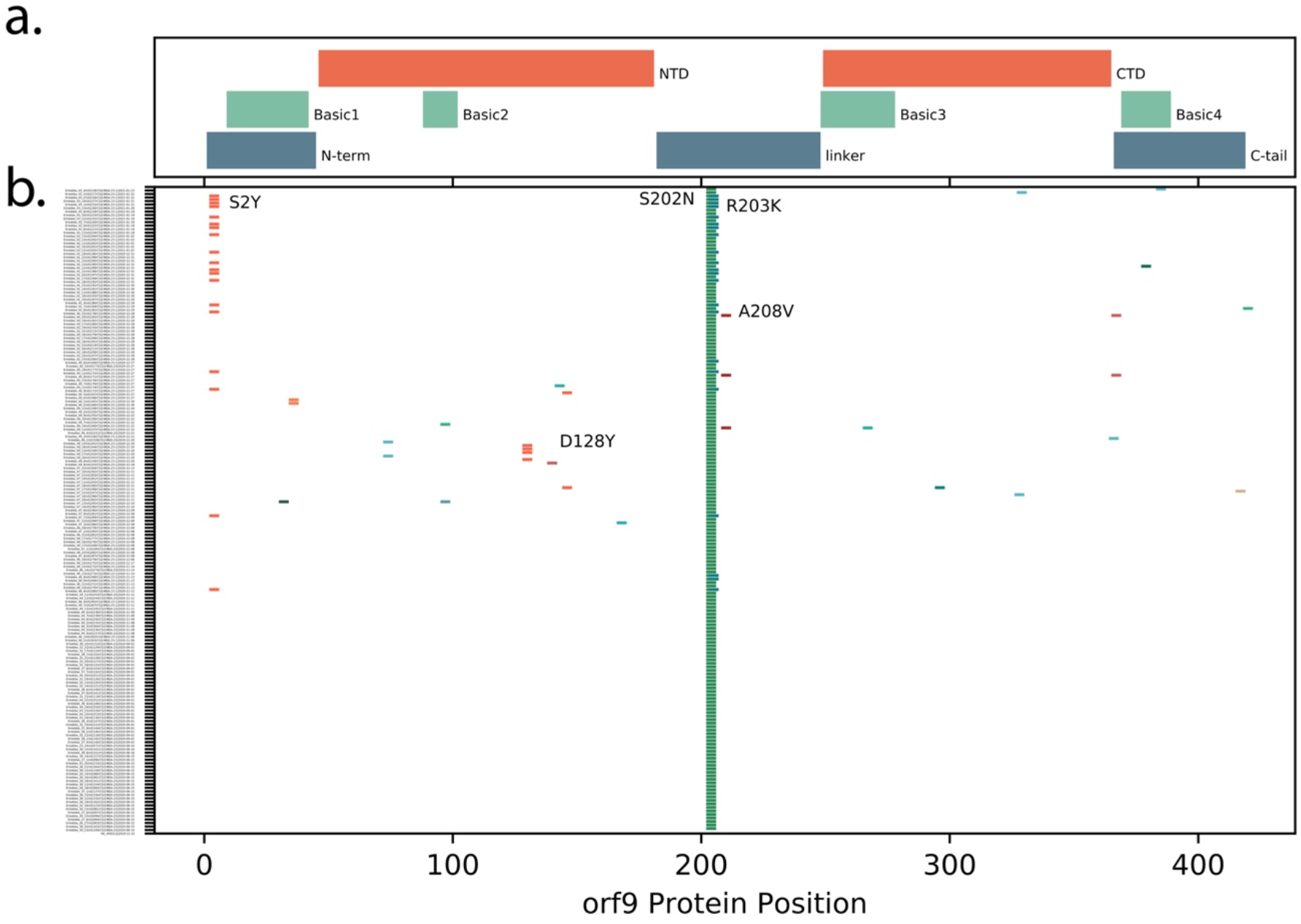
Changes in A.23/A.23.1 ORF9 protein. The encoded ORF9 protein from all Ugandan A.23 and A.23.1 genomes gather, aligned and compared to the ORF9 protein from GenBank NC_045512.2. **Panel a**: The locations of important ORF9 protein features are indicated based on the analysis of ORF9 from Chang et al.(41). N-term: amino-terminal extension, NTD: amino-terminal domain, linker: linker region between the NTD and CTD, CTD: carboxy-terminal domain, C-tail: carboxy-terminal extension, Regions with “Basic” indicate the 4 regions enriched in positively charged amino acids. **Panel b:** Each line represents the encoded ORF9 protein sequence from a single genome, ordered by date of samples collection (bottom earliest, top most recent). Markers indicating the positions of amino acid (aa) differences from the reference strain, changes observed in multiple genomes are annotated with the annotation (original aa position new aa).

## Notes

### Competing Interest Statement

The authors have declared no competing interest.

### Funding Statement

The SARS-CoV2 diagnostic and sequencing award is jointly funded by the UK Medical Research Council (MRC) and the UK Department for International Development (DFID) under the MRC/DFID Concordat agreement (grant agreement number NC_PC_19060) and is also part of the EDCTP2 programme supported by the European Union. The UMIC high performance computer was supported by MRC (grant number MC_EX_MR/L016273/1) to PK. A.R. acknowledges the support of the Wellcome Trust (Collaborators Award 206298/Z/17/Z ARTIC network) and the European Research Council (grant agreement no. 725422 ReservoirDOCS). The study is additionally funded by the Wellcome, DFID Wellcome Epidemic Preparedness Coronavirus (grant agreement number 220977/Z/20/Z) awarded to MC.

### Author Declarations

This study was approved by the Uganda Virus Research Institute- Research and Ethics Committee (UVRI-REC Federalwide Assurance [FWA] FWA No. 00001354, study reference. GC/127/20/04/771) and by the Uganda National Council for Science and Technology, reference number HS936ES.

## References

1. Edward C. Holmes, Yong-Zhen Zhang Ec. Initial genome release of novel coronavirus. Virological.org [Internet]. 2020 [cited 2021 Jan 24]; Available from: http://virological.org/t/319

2. Li Q, Guan X, Wu P, Wang X, Zhou L, Tong Y, et al. Early Transmission Dynamics in Wuhan, China, of Novel Coronavirus–Infected Pneumonia. N Engl J Med. 2020 Jan 29;EJMoa2001316.

3. Yang X, Yu Y, Xu J, Shu H, Xia J, Liu H, et al. Clinical course and outcomes of critically ill patients with SARS-CoV-2 pneumonia in Wuhan, China: a single-centered, retrospective, observational study. Lancet Respir Med. 2020 Feb;S2213260020300795.

4. Rambaut A, Holmes EC, O’Toole Á, Hill V, McCrone JT, Ruis C, et al. A dynamic nomenclature proposal for SARS-CoV-2 lineages to assist genomic epidemiology. Nat Microbiol. 2020 Nov;5(11):1403–7.

5. Volz E, Mishra S, Chand M, Barrett JC, Johnson R, Geidelberg L, et al. Transmission of SARS-CoV-2 Lineage B.1.1.7 in England: Insights from linking epidemiological and genetic data [Internet]. Infectious Diseases (except HIV/AIDS); 2021 Jan [cited 2021 Jan 29]. Available from: http://medrxiv.org/lookup/doi/10.1101/2020.12.30.20249034

6. Tegally H, Wilkinson E, Giovanetti M, Iranzadeh A, Fonseca V, Giandhari J, et al. Emergence and rapid spread of a new severe acute respiratory syndrome-related coronavirus 2 (SARS-CoV-2) lineage with multiple spike mutations in South Africa [Internet]. Epidemiology; 2020 Dec [cited 2021 Jan 6]. Available from: http://medrxiv.org/lookup/doi/10.1101/2020.12.21.20248640

7. Voloch CM, da Silva Francisco R, de Almeida Lgp, Cardoso CC, Brustolini OJ, Gerber AL, et al. Genomic characterization of a novel SARS-CoV-2 lineage from Rio de Janeiro, Brazil. J Virol. 2021 Mar 1;

8. Bugembe DL, Kayiwa J, Phan MVT, Tushabe P, Balinandi S, Dhaala B, et al. Main Routes of Entry and Genomic Diversity of SARS-CoV-2, Uganda. Emerg Infect Dis. 2020 Oct;26(10):2411–5.

9. Hadfield J, Megill C, Bell SM, Huddleston J, Potter B, Callender C, et al. Nextstrain: real-time tracking of pathogen evolution. Kelso J, editor. Bioinformatics. 2018 Dec 1;34(23):4121–3.

10. Githinji G, de Laurent ZR, Mohammed KS, Omuoyo DO, Macharia PM, Morobe JM, et al. Tracking the introduction and spread of SARS-CoV-2 in coastal Kenya [Internet]. Epidemiology; 2020 Oct [cited 2020 Dec 7]. Available from: http://medrxiv.org/lookup/doi/10.1101/2020.10.05.20206730

11. Page AJ, Mather AE, Le Viet T, Meader EJ, Alikhan N-FJ, Kay GL, et al. Large scale sequencing of SARS-CoV-2 genomes from one region allows detailed epidemiology and enables local outbreak management [Internet]. Epidemiology; 2020 Sep [cited 2020 Oct 9]. Available from: http://medrxiv.org/lookup/doi/10.1101/2020.09.28.20201475

12. Filipe ADS, Shepherd J, Williams T, Hughes J, Aranday-Cortes E, Asamaphan P, et al. Genomic epidemiology of SARS-CoV-2 spread in Scotland highlights the role of European travel in COVID-19 emergence [Internet]. Infectious Diseases (except HIV/AIDS); 2020 Jun [cited 2020 Dec 14]. Available from: http://medrxiv.org/lookup/doi/10.1101/2020.06.08.20124834

13. Daily Monitor. Amuru prison closed as 153 test positive for Covid-19. 2020 Aug 23; Available from: https://www.monitor.co.ug/uganda/news/national/amuru-prison-closed-as-153-test-positive-for-covid-19-1924660

14. Penelope Nankunda. COVID-19: Uganda registers 318 new cases in a single day. MSN [Internet]. 2020 Aug 22; Available from: https://www.msn.com/en-xl/news/other/covid-19-uganda-registers-318-new-cases-in-a-single-day/ar-BB18gprA

15. Li Q, Wu J, Nie J, Zhang L, Hao H, Liu S, et al. The Impact of Mutations in SARS-CoV-2 Spike on Viral Infectivity and Antigenicity. Cell. 2020 Sep 3;182(5):1284-1294.e9.

16. Nguyen HT, Zhang S, Wang Q, Anang S, Wang J, Ding H, et al. Spike glycoprotein and host cell determinants of SARS-CoV-2 entry and cytopathic effects. J Virol. 2020 Dec 11;

17. Gobeil SM-C, Janowska K, McDowell S, Mansouri K, Parks R, Manne K, et al. D614G Mutation Alters SARS-CoV-2 Spike Conformation and Enhances Protease Cleavage at the S1/S2 Junction. Cell Rep. 2021 Jan 12;34(2):108630.

18. Volz E, Hill V, McCrone JT, Price A, Jorgensen D, O’Toole Á, et al. Evaluating the Effects of SARS-CoV-2 Spike Mutation D614G on Transmissibility and Pathogenicity. Cell. 2020 Nov;S0092867420315373.

19. Áine O’Toole et al. Phylogenetic Assignment of Named Global Outbreak LINeages (PANGOLIN). 2020; Available from: https://github.com/cov-lineages/pangolin

20. Hoffmann M, Kleine-Weber H, Pöhlmann S. A Multibasic Cleavage Site in the Spike Protein of SARS-CoV-2 Is Essential for Infection of Human Lung Cells. Mol Cell. 2020 May;78(4):779-784.e5.

21. Áine O’Toole et al. B.1.1.7 report 2021-02-05. 2021; Available from: https://cov-lineages.org/global_report_B.1.1.7.html

22. Áine O’Toole, JT McCrone, Verity Hill and Andrew Rambaut. Pangolin COVID-19 Lineage Assigner. Available from: https://pangolin.cog-uk.io/

23. Rambaut A, Holmes EC, Hill V, O’Toole Á, McCrone J, Ruis C, et al. A dynamic nomenclature proposal for SARS-CoV-2 to assist genomic epidemiology [Internet]. Microbiology; 2020 Apr [cited 2020 Apr 27]. Available from: http://biorxiv.org/lookup/doi/10.1101/2020.04.17.046086

24. Phan MVT, Ngo Tri T, Hong Anh P, Baker S, Kellam P, Cotten M. Identification and characterization of Coronaviridae genomes from Vietnamese bats and rats based on conserved protein domains. Virus Evol [Internet]. 2018 Jul 1 [cited 2020 Jan 12];4(2). Available from: https://academic.oup.com/ve/article/doi/10.1093/ve/vey035/5250438

25. Edgar RC. Search and clustering orders of magnitude faster than BLAST. Bioinformatics. 2010 Oct 1;26(19):2460–1.

26. Katoh K, Standley DM. MAFFT Multiple Sequence Alignment Software Version 7: Improvements in Performance and Usability. Mol Biol Evol. 2013 Apr 1;30(4):772–80.

27. Eddy SR. Accelerated Profile HMM Searches. PLOS Comput Biol. 2011 Oct 20;7(10):e1002195.

28. Tegally H, Wilkinson E, Giovanetti M, Iranzadeh A, Fonseca V, Giandhari J, et al. Emergence of a SARS-CoV-2 variant of concern with mutations in spike glycoprotein. Nature [Internet]. 2021 Mar 9 [cited 2021 Mar 12]; Available from: http://www.nature.com/articles/s41586-021-03402-9

29. Benvenuto D, Angeletti S, Giovanetti M, Bianchi M, Pascarella S, Cauda R, et al. Evolutionary analysis of SARS-CoV-2: how mutation of Non-Structural Protein 6 (NSP6) could affect viral autophagy. J Infect. 2020 Jul;81(1):e24–7.

30. Voloch CM, Silva F R da, de Almeida LGP, Cardoso CC, Brustolini OJ, Gerber AL, et al. Genomic characterization of a novel SARS-CoV-2 lineage from Rio de Janeiro, Brazil [Internet]. Genetic and Genomic Medicine; 2020 Dec [cited 2021 Jan 30]. Available from: http://medrxiv.org/lookup/doi/10.1101/2020.12.23.20248598

31. Su YCF, Anderson DE, Young BE, Linster M, Zhu F, Jayakumar J, et al. Discovery and Genomic Characterization of a 382-Nucleotide Deletion in ORF7b and ORF8 during the Early Evolution of SARS-CoV-2. Schultz-Cherry S, editor. mBio. 2020 Jul 21;11(4):e01610–20, /mbio/11/4/mBio.01610-20.atom.

32. The Chinese SARS Molecular Epidemiology Consortium. Molecular Evolution of the SARS Coronavirus During the Course of the SARS Epidemic in China. Science. 2004 Mar 12;303(5664):1666–9.

33. Cotten M, Bugembe DL, Kaleebu P, Phan MVT. Alternate primers for whole-genome SARS-CoV-2 sequencing [Internet]. Genomics; 2020 Oct [cited 2020 Nov 30]. Available from: http://biorxiv.org/lookup/doi/10.1101/2020.10.12.335513

34. Li H. Minimap2: pairwise alignment for nucleotide sequences. Birol I, editor. Bioinformatics. 2018 Sep 15;34(18):3094–100.

35. Larsson A. AliView: a fast and lightweight alignment viewer and editor for large datasets. Bioinformatics. 2014 Nov 15;30(22):3276–8.

36. Kozlov AM, Darriba D, Flouri T, Morel B, Stamatakis A. RAxML-NG: a fast, scalable and user-friendly tool for maximum likelihood phylogenetic inference. Bioinforma Oxf Engl. 2019 Nov 1;35(21):4453–5.

37. Darriba D, Posada D, Kozlov AM, Stamatakis A, Morel B, Flouri T. ModelTest-NG: A New and Scalable Tool for the Selection of DNA and Protein Evolutionary Models. Mol Biol Evol. 2020 Jan 1;37(1):291–4.

38. Rambaut A. FigTree http://tree.bio.ed.ac.uk/software/figtree. 2019;

39. Josh B. Singer, Gifford R, Cotten M, Robertson DL. CoV-GLUE project. 2020; Available from: http://cov-glue.cvr.gla.ac.uk/

40. Flower TG, Buffalo CZ, Hooy RM, Allaire M, Ren X, Hurley JH. Structure of SARS-CoV-2 ORF8, a rapidly evolving coronavirus protein implicated in immune evasion [Internet]. Biophysics; 2020 Aug [cited 2021 Mar 14]. Available from: http://biorxiv.org/lookup/doi/10.1101/2020.08.27.270637

41. Chang C, Hou M-H, Chang C-F, Hsiao C-D, Huang T. The SARS coronavirus nucleocapsid protein – Forms and functions. Antiviral Res. 2014 Mar;103:39–50.

